# Explicit Motor Imaging Abilities are Similar in Complex Regional Pain Syndrome, Chronic Limb Pain and Healthy Individuals: a cross sectional study

**DOI:** 10.1101/2023.04.02.23288051

**Authors:** G. Cohen-Aknine, A.F. Homs, D. Mottet, T. Mura, F. Jedryka, A. Dupeyron

**Affiliations:** EuroMov Digital Health in Motion, Univ Montpellier, IMT Mines Ales, CHU Nimes, Nimes, France; EuroMov Digital Health in Motion, Univ Montpellier, IMT Mines Ales, Univ Montpellier, France; Department of Biostatistics, Epidemiology, Public Health and Innovation in Methodology (BESPIM), CHU Nimes, Nimes, Univ Montpellier, France; Department of Pain Management, CHU Nimes, Univ Montpellier, Nimes, France

## Abstract

**Background:** Complex regional pain syndrome (CRPS) is a chronic pain condition characterised by peripheral and central sensory and motor dysfunction. Implicit motor imagery is known to be impaired in these patients, but evidence is still lacking for explicit motor imagery. Using a self-rated questionnaire, this study aims to compare explicit motor imagery abilities between individuals with CRPS, with chronic limb pain (CLP) and healthy controls and also examine differences between affected and unaffected limbs.

**Methods:** In this single-centre observational study, 123 participants were recruited (CRPS = 40, chronic limb pain, CLP = 40, and healthy individuals = 43). Participants completed the Movement Imagery Questionnaire - Revised Second (MIQ-RS) once for each body side. The total MIQ-RS score, and the kinesthetic and visual subscores were compared between groups and between the affected and unaffected sides.

**Results:** The MIQ-RS revealed no significant differences in explicit motor imagery abilities, neither between groups nor between the affected and unaffected side. Null Hypothesis Bayesian Testing on kinesthetic motor imagery abilities indicated a sevenfold likelihood of no differences between groups and a more than fivefold likelihood of no differences between sides.

**Conclusion:** CRPS and chronic limb pain individuals showed preserved explicit motor imagery abilities, notably on the pain side. The preservation of these abilities supports the recommendation of mental imagery therapy to improve motor function and relieve pain in chronic pain patients.

## INTRODUCTION

Complex regional pain syndrome (CRPS) is a rare, debilitating condition that primarily affects a single extremity and is characterised by pain that is disproportionate to the original injury (M. C. Ferraro et al., 2024; Goebel et al., 2019). The International Association for the Study of Pain (IASP) classifies CRPS as chronic primary pain in the ICD-11 and suggests that it may also meet the criteria for non-disabling pain (Goebel et al., 2021; Harden et al., 2010a; Kosek et al., 2021). The pathophysiology of CRPS involves complex interactions between immune-mediated inflammatory responses, vasomotor changes, genetic factors, psychological components and changes in the nervous system. These changes cause sensory, motor, autonomic and trophic dysfunctions (Birklein et al., 2018; M. C. Ferraro et al., 2024) such as allodynia or hyperalgesia, with 63% of patients experiencing a reduction in active movement (Ott & Maihöfner, 2018). Sensory and motor dysfunction are associated with cortical changes, referred to as maladaptive plasticity (Ma et al., 2022; Shokouhi et al., 2018; Zangrandi et al., 2021).

Engaging patients in active approaches improves motor recovery and promotes brain plasticity (M. Ferraro et al., 2023; Harden et al., 2022; Smart et al., 2022). This has motivated the use of motor imagery therapy, alone or in combination with other therapies, to activate motor neural networks with reduced pain associated with physical movement (Lotze & Moseley, 2022a). Motor imagery (MI) is a dynamic state in which individuals mentally simulate specific actions or gestures without actually performing the movement (Decety, 1996; Moran et al., 2012). Motor imagery training has shown promise in improving function and reducing pain by activating neural pathways similar to those used during actual movement execution (M. Ferraro et al., 2023; Hardwick et al., 2018; Ríos-León et al., 2024).

Despite various theoretical models proposing explanations for the motor imagery phenomenon, such as the motor simulation theory and the effect imagery model, there is no consensus on a definitive explanation (Hurst & Boe, 2022; Solomon et al., 2022)

These therapies require patients to imagine their affected side in different modalities, and assessment methods are heterogeneous, ranging from implicit tasks (lateral judgement) to explicit tasks (self-report questionnaires, mental chronometry) detailing visual and kinesthetic perspectives (Chepurova et al., 2022; Guillot & Collet, 2005). Previous studies have shown that individuals with CRPS have significant deficits in explicit MI compared to healthy controls and their unaffected side (Breckenridge et al., 2019; Ravat et al., 2020). Furthermore, a study by La Touche et al. (La Touche et al., 2019) showed that explicit MI difficulties are more pronounced in individuals with chronic low back pain than in asymptomatic controls. However, explicit MI abilities in people with CRPS are poorly understood.

Consistent with the observed impairments in implicit motor imagery in patients with chronic limb pain compared to healthy individuals and their unaffected limbs, we hypothesize that CRPS patients will show specific deficits in explicit motor imagery abilities. This study compares explicit motor imagery between CRPS patients, chronic limb pain patients, healthy controls, and between affected and unaffected limbs.

## MATERIAL AND METHODS

### Design

This was a prospective, single-centre, cross-sectional study conducted at the University Hospital of Nîmes (France). The study procedures complied with the ethical standards of the competent committee for human experimentation (local ethics committee 2020-A02281-38 designated “Comité de Protection des Personnes, Sud Méditérrannée IV” on 8/12/2020) and the Helsinki Declaration of 2013. The study protocol was registered on clinicaltrials.org on 11/01/2021 (NCT04703348). All individuals received an information letter and informed consent was obtained from all individuals.

### Participants and setting

Individuals with CRPS were recruited at the Department of Pain Medicine (CHU Nîmes, France) between January 2021 and October 2022. Pain specialists recruited patients based on the verification of the presence of the Budapest and eligibility criteria. Patients with CRPS were affected in either the upper or lower limb and on the dominant or non-dominant side. Healthy individuals were recruited through a poster campaign among hospital staff. Patients with chronic limb pain (CLP) were recruited and diagnosed at the Department of Physical Medicine and Rehabilitation by specialists in physical medicine and rehabilitation. They were included in the study if they had experienced limb pain for more than three months due to conditions such as musculoskeletal disorders, chronic post-traumatic pain, or post-operative pain, regardless of the underlying cause.

Inclusion criteria were: Age over 18 years, less than 150 minutes of moderate-to-vigorous physical activity per week, and education up to high school graduation or equivalent.

Patients with CRPS had to meet the diagnostic criteria set out in the Budapest criteria (Harden et al., 2010b; Mesaroli et al., 2021). Both CRPS patients and those with chronic limb pain needed to have experienced pain for at least three months. In addition, individuals were excluded from the study if they met any of the following criteria CRPS secondary to stroke, stellate block performed within three weeks prior to the interview, presence of a central neurological disorder, diagnosis of chronic fibromyalgia or low back pain, pregnancy, postpartum or lactation, visual impairment that interfered with the use of the MIQ-RS questionnaire, history of limb amputation, previous experience with motor imagery practice, or psychiatric illness.

To characterise the population age, sex, body mass index (BMI), upper and lower dominant limb, education level, pain duration and physical activity level were recorded.

### Protocol

Prior to assessing the eligibility criteria and obtaining informed consent from the physician, individuals were asked to complete the Movement Imagery Questionnaire - Revised, Second Edition (MIQ-RS) during the consultation. Age, weight, height, pain duration and limb dominance (upper and lower) were self-reported. Patients were also asked to report their level of physical activity on a three-point scale (less than 1 hour, between 1 and 1.5 hours, and more than 1.5 hours of moderate to vigorous activity per week) and their level of education on a six-point scale (from high school to PhD and beyond). The Movement Imagery Questionnaire-Revised, Second Edition (MIQ-RS) was chosen for several reasons: its suitability for patients with motor limitations (Gregg et al., 2010), its ability to measure lateralised imagery scores (comparing left and right sides), its validation in French (Loison et al., 2013), and its acceptable reliability and reproducibility (Butler et al., 2012). Furthermore, this questionnaire has been shown to correlate visual and kinesthetic scores with fMRI signals in stroke patients (Confalonieri et al., 2012), although it has not yet been used in patients with CRPS.

However, due to the Covid-19 pandemic, some individuals (60%) completed the questionnaire via videoconference with an investigator. The questionnaires were audio-recorded on REDCap© (online questionnaire) (Floridou et al., 2022). Pain individuals completed a self-rated questionnaire twice, starting with the right side and then answering the left side, pausing if necessary. The session was administered in a single session, with no follow-up. The expected heterogeneity in dominance, laterality and upper or lower limb affected did not allow for randomisation.

### Outcome measures

The MIQ-RS is a validated self-rated questionnaire for the assessment of explicit motor imagery (Butler et al., 2012; Gregg et al., 2010). It is a 14-item therapist-administered questionnaire in which patients first perform a movement, e.g. raising the knee, followed by visual and then kinesthetic motor imagery. Patients rate their abilities on a 7-point Likert scale, ranging from “very easy to see/feel” (1 point) to “very hard to see/feel” (7 points). The MIQ-RS offers two methods for scoring, as documented in the literature: the first method involves the calculation of a total score and two subscores for Kinesthetic Motor Imagery (KMI) and Visual Motor Imagery (VMI). The total possible score is 98, with each of the subscores (KMI and VMI) having a maximum of 49 points (Rimbert et al., 2019). Alternatively, the score can be derived by taking the mean of the responses for the total score and the two subscores on a 7-point Likert scale. There is no official cutoff point for assessing explicit motor imagery abilities. However, based on the systematic review by McInnes et al. (2016), explicit motor imagery abilities were categorized into three levels: unable (scoring less than 48 out of 98), impaired (scoring between 49 and 73 out of 98), and normal (scoring more than 74 out of 98).

The primary outcome of the study was the categorisation of motor imagery (MI) abilities for the three groups of participants. Secondary outcomes included differences in total and subscores of the MIQ-RS between the groups and differences in scores on the unaffected and affected sides within the CRPS and CLP groups.

No published results for this population were available for sample size calculation. Based on previous research using the MIQ-RS in patients with chronic conditions such as chronic low back pain, where La Touche et al. (La Touche et al., 2019) found an effect size of 0.57 between groups and aiming for 95% power with a beta risk of 0.80 to compare three groups, GPower 3.1.9.7 was used to calculate the required sample size. The initial power calculation suggested 33 individuals per group. However, to account for a potential 20% exclusion due to eligibility criteria, the number was adjusted to 40 individuals per group. This adjustment results in a total of 120 participants required for the study.

### Data analysis

The softwares JASP © and R Studio © were used to perform the statistical analyses.

First, a frequentist statistical approach was used with significance set for a two-tailed α level of 0.05. As data were not distributed normally, we used non-parametric signed-rank tests and report median and interquartile range (IQR) with a 95% confidence interval. To assess the effect of group (CRPS vs. CLP vs. healthy) on motor imagery scores, we performed a Kruskal-Wallis test. To assess the effect of pain on motor imagery scores across limbs (unaffected vs. affected), we performed a Wilcoxon signed-rank test. Finally, we assessed the dispersion of scores between groups using a coefficient of variation.

Secondly, we used a Null Hypothesis Bayesian Testing (NHBT) approach for the assessment of evidence of a lack of difference (Kruschke, 2021; van Doorn et al., 2021). Specifically, we calculated the Bayes factor (BF01), which quantifies the likelihood of the null hypothesis versus the alternative hypothesis (for example, BF01 = 7.13 indicates that, given the data, no difference is 7.13 times more likely than a difference). A Bayes factor between 5 and 10 indicates moderate evidence in favour of the null hypothesis (Quintana & Williams, 2018). Levene’s method was used to measure the equality of variances between the groups and sides. This analysis was performed using JASP © software with a prior in favour of differences between groups according to our hypothesis (van Doorn et al., 2021).

## RESULTS

We screened 129 participants for inclusion, and 123 were included after exclusion and age matching (40 participants in the CRPS and CLP group and 43 participants in the healthy group). The main characteristics of the participants are described in Table 1 (further demographic details are provided in Appendix S1).

**Table 1:**
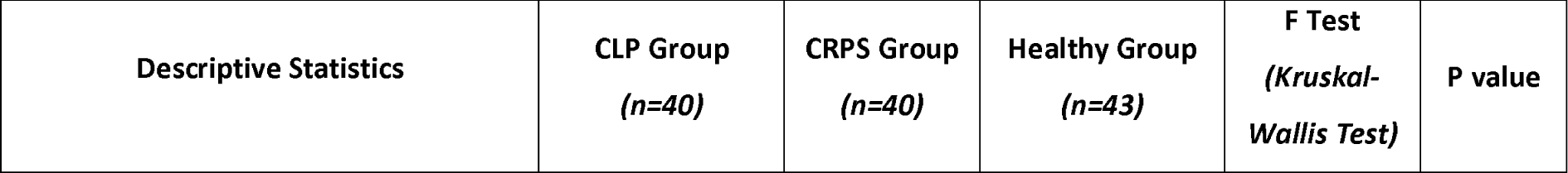
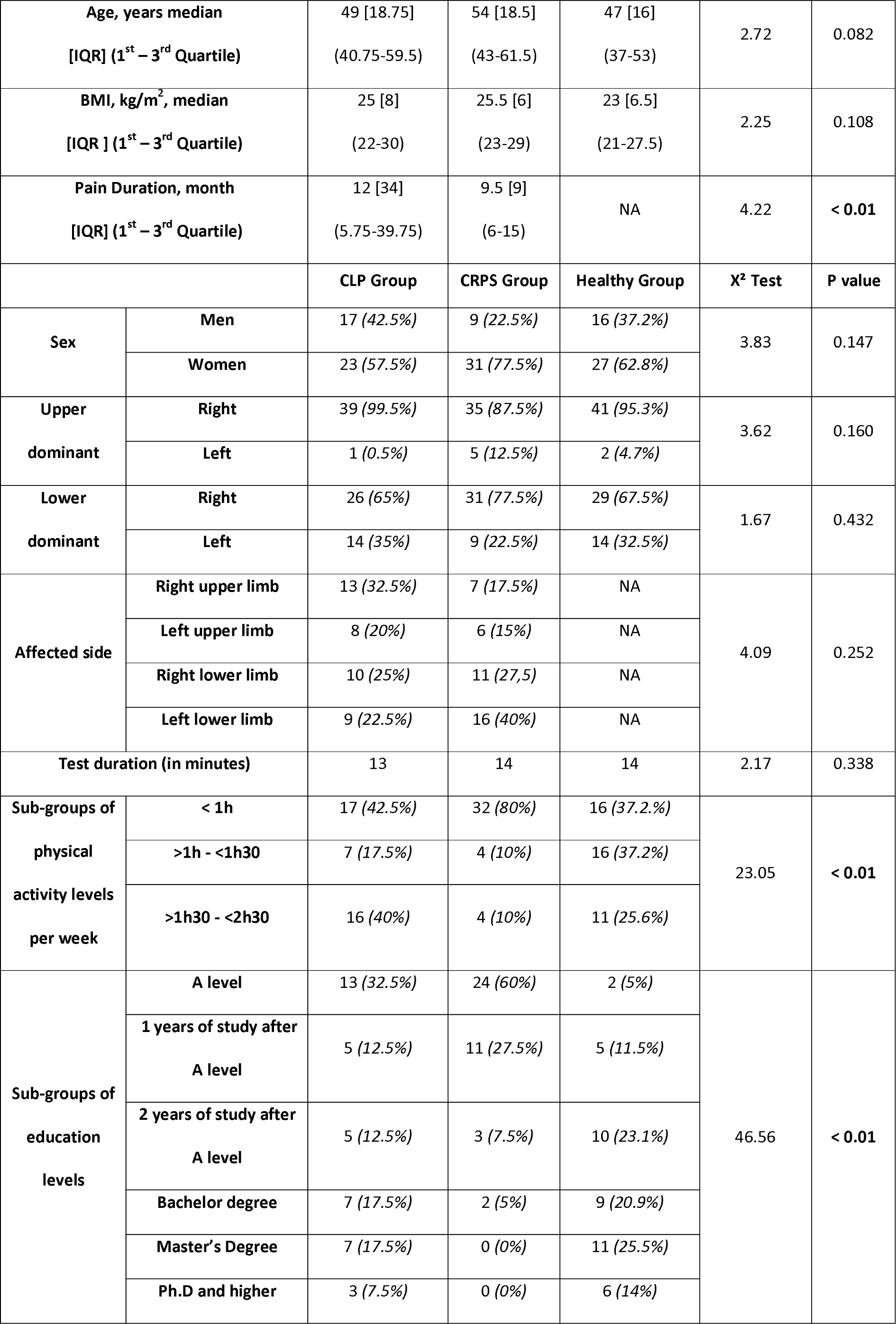
Participant demographic characteristics. Data are presented as medians for continuous variables and as numbers and percentages for categorical variables in each group.

| Descriptive Statistics | CLP Group<br>(n=40) | CRPS Group<br>(n=40) | Healthy Group<br>(n=43) | F Test<br>(Kruskal-Wallis Test) | P value |
| --- | --- | --- | --- | --- | --- |

|  |  |  |  |  |  |  |
| --- | --- | --- | --- | --- | --- | --- |
| **Age, years median**<br>**[IQR] (1<sup>st</sup> – 3<sup>rd</sup> Quartile)** |  | 49 [18.75]<br>(40.75-59.5) | 54 [18.5]<br>(43-61.5) | 47 [16]<br>(37-53) | 2.72 | 0.082 |
| **BMI, kg/m<sup>2</sup>, median**<br>**[IQR ] (1<sup>st</sup> – 3<sup>rd</sup> Quartile)** |  | 25 [8]<br>(22-30) | 25.5 [6]<br>(23-29) | 23 [6.5]<br>(21-27.5) | 2.25 | 0.108 |
| **Pain Duration, month**<br>**[IQR] (1<sup>st</sup> – 3<sup>rd</sup> Quartile)** |  | 12 [34]<br>(5.75-39.75) | 9.5 [9]<br>(6-15) | NA | 4.22 | **< 0.01** |
|  |  | **CLP Group** | **CRPS Group** | **Healthy Group** | **X<sup>2</sup> Test** | **P value** |
| **Sex** | **Men** | 17 (42.5%) | 9 (22.5%) | 16 (37.2%) | 3.83 | 0.147 |
|  | **Women** | 23 (57.5%) | 31 (77.5%) | 27 (62.8%) |  |  |
| **Upper dominant** | **Right** | 39 (99.5%) | 35 (87.5%) | 41 (95.3%) | 3.62 | 0.160 |
|  | **Left** | 1 (0.5%) | 5 (12.5%) | 2 (4.7%) |  |  |
| **Lower dominant** | **Right** | 26 (65%) | 31 (77.5%) | 29 (67.5%) | 1.67 | 0.432 |
|  | **Left** | 14 (35%) | 9 (22.5%) | 14 (32.5%) |  |  |
| **Affected side** | **Right upper limb** | 13 (32.5%) | 7 (17.5%) | NA | 4.09 | 0.252 |
|  | **Left upper limb** | 8 (20%) | 6 (15%) | NA |  |  |
|  | **Right lower limb** | 10 (25%) | 11 (27,5) | NA |  |  |
|  | **Left lower limb** | 9 (22.5%) | 16 (40%) | NA |  |  |
| **Test duration (in minutes)** |  | 13 | 14 | 14 | 2.17 | 0.338 |
| **Sub-groups of physical activity levels per week** | **< 1h** | 17 (42.5%) | 32 (80%) | 16 (37.2%) | 23.05 | **< 0.01** |
|  | **>1h - <1h30** | 7 (17.5%) | 4 (10%) | 16 (37.2%) |  |  |
|  | **>1h30 - <2h30** | 16 (40%) | 4 (10%) | 11 (25.6%) |  |  |
| **Sub-groups of education levels** | **A level** | 13 (32.5%) | 24 (60%) | 2 (5%) | 46.56 | **< 0.01** |
|  | **1 years of study after A level** | 5 (12.5%) | 11 (27.5%) | 5 (11.5%) |  |  |
|  | **2 years of study after A level** | 5 (12.5%) | 3 (7.5%) | 10 (23.1%) |  |  |
|  | **Bachelor degree** | 7 (17.5%) | 2 (5%) | 9 (20.9%) |  |  |
|  | **Master's Degree** | 7 (17.5%) | 0 (0%) | 11 (25.5%) |  |  |
|  | **Ph.D and higher** | 3 (7.5%) | 0 (0%) | 6 (14%) |  |  |

### Identification of Motor Imagery abilities

The MIQ-RS total scores for participant with complex regional pain syndrome, chronic limb pain and matched healthy participants showed high heterogeneity. This variability meant that groups of participants could not be categorised as having no, impaired or normal motor imagery ability based on the MIQ-RS total scores for both the healthy and painful side (Fig. 1).

**Figure 1:**
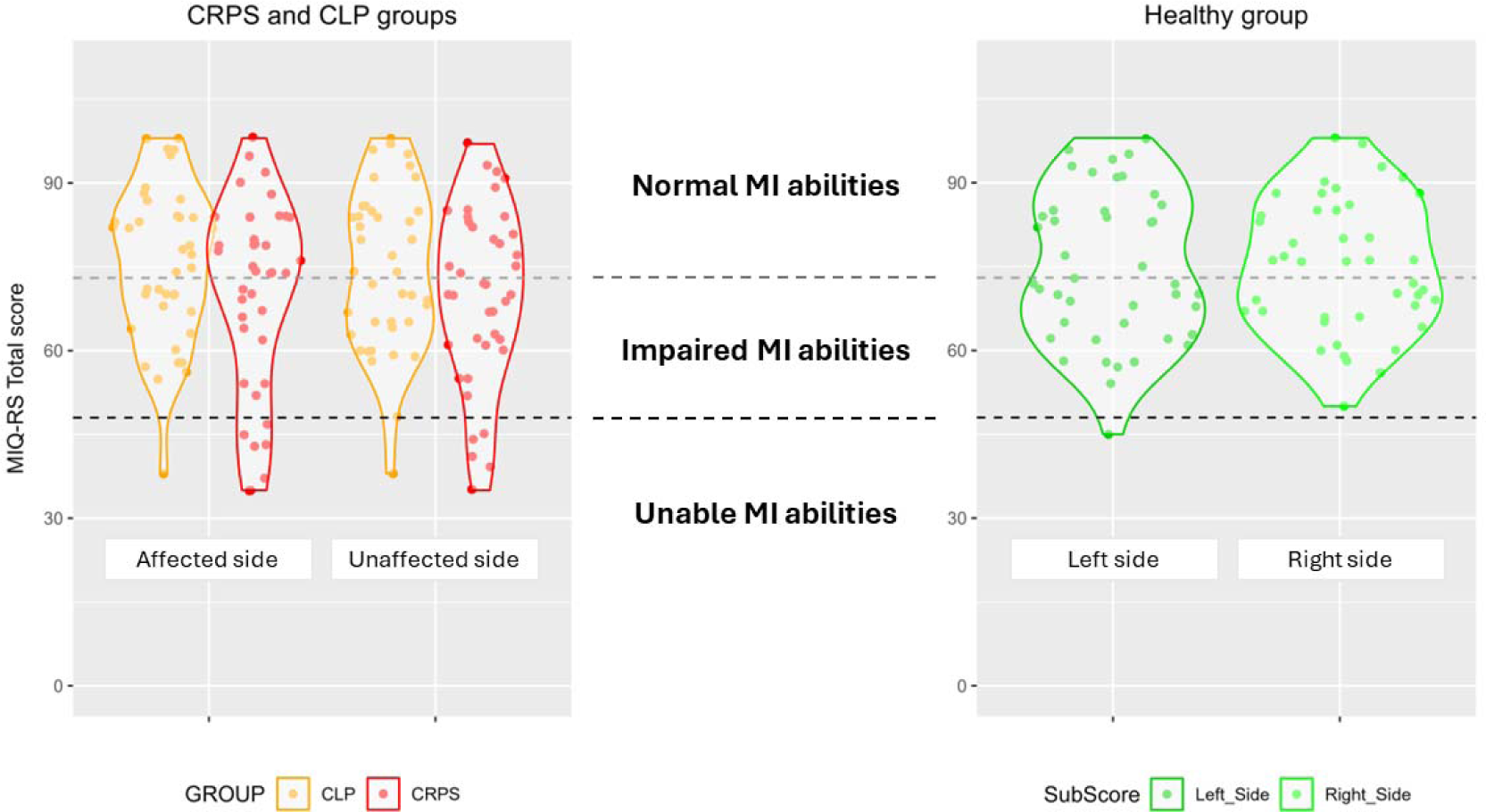
Violin plot of categorisation of total MIQ-RS scores between complex regional pain syndrome, chronic limb pain and healthy groups.

Indeed, if we apply the classification proposed by McInnes et al. (McInnes et al., 2016), which was developed based on participants with brain lesions, it appears that all groups, including those with complex regional pain syndrome, chronic limb pain and healthy participants, show impairments (between 49 and 73) in explicit motor imagery even in unaffected side (Appendix S1, Table S2). The median values of the MIQ-RS Total scores for the affected side and the unaffected side are shown in Appendix S1 (Table S2).

### Between-Individuals coefficient of variation

The coefficient of variation between individuals for the CRPS group is 24.53% for the affected side and 22.52% for the unaffected side. For the CLP group, the coefficient of variation is 18.13% for the affected side and 19.05% for the unaffected side. For the healthy group, the coefficient of variation is 15.84% for the right side and 17.85% for the left side. These results show a high dispersion of results, especially for the CRPS group, but with consistency between the affected and unaffected side for the pain groups.

### Comparison of MI abilities between groups

There were no statistical differences between the three groups for the MIQ-RS total mean score (H(2) = 1.795, p = 0.408, n² = -0.002) (Figure 2), the kinesthetic mean subscore (H(2) = 0.936, p = 0.626, n² = -0.009), or the visual mean subscore (H(2) = 4.175, p = 0.124, n² = 0.018) (Appendix S1, Table S3).

**Figure 2:**
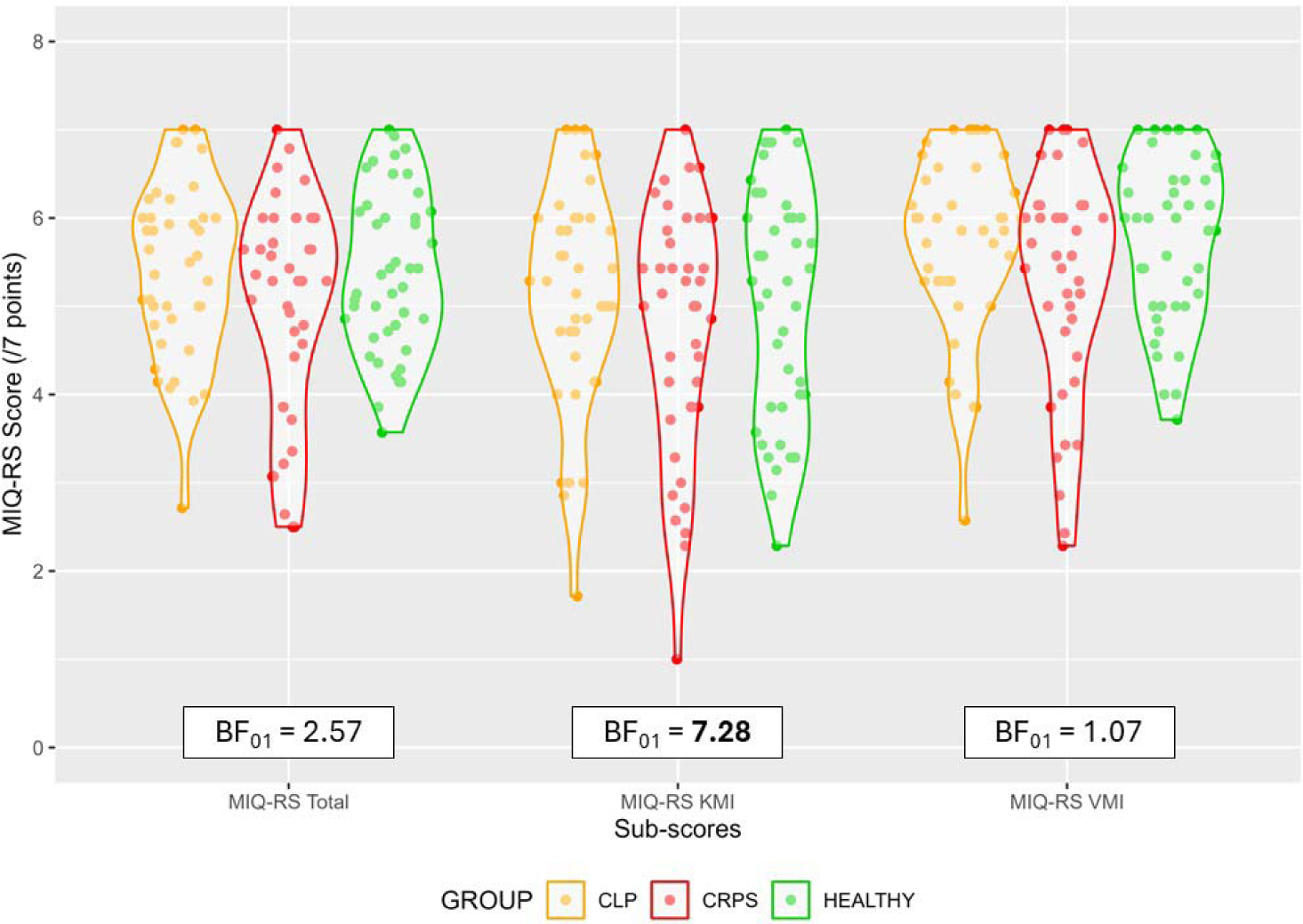
Violin plots of MIQ-RS mean scores between complex regional pain syndrome, chronic limb pain and healthy groups with Bayesian null hypothesis tests (BF01) between groups.

### Comparison of MI abilities between the affected and unaffected sides

#### Complex regional pain syndrome Group analysis

There was no statistical difference between the affected side and the unaffected side for the MIQ-RS total mean score (W = 347.500, p = 0.826, r = 0.148), kinesthetic mean subscore (W = 377.000, p = 0.706, r = 0.086) and visual mean subscore (W = 272.500, p = 0.0881, r = 0.0187).The results are summarised in Appendix S1 (Table S4).

#### Chronic limb pain group analysis

There was also no statistical difference between the affected side and the unaffected side for the MIQ-RS total mean score (W = 343.500, p = 0.910, r = 0.148), kinesthetic mean subscore (W = 372.500, p = 0.983, r = 0.334) and visual mean subscores (W = 317.500, p = 0.514, r = 0.006) in the CLP group (Appendix S1, Table S4).

The complete inferential statistical analysis can be found in the Supplementary Information document (Appendix S2 in html).

### Bayesian Null Hypothesis Testing

Frequentist analysis showed no significant differences between the groups or between the affected and unaffected sides. Therefore, a Bayesian null hypothesis testing approach was used to draw conclusions about the null hypothesis and group similarities (Kruschke, 2021).

Thus, we confirmed the absence of between-group differences in explicit motor imagery abilities with moderate evidence for the null hypothesis (BF_01_ > 5), specifically in Kinesthetic Motor Imagery (KMI) with a Bayesian factor of 7.28, as detailed in Appendix S1 (Table S5).

We also confirmed the absence of differences in explicit motor imagery abilities between the affected and unaffected side in both the CRPS and CLP groups, with moderate evidence for the null hypothesis (BF_01_ > 5). Details are provided in Appendix S1 (Table S6).

This indicates that chronic pain does not affect explicit motor imagery abilities as assessed by the MIQ-RS (Fig. 2).

In simpler terms, our analysis suggests that there is a 7-fold probability that there are no differences in kinesthetic motor imagery abilities between the groups. Similarly, when comparing motor imagery (both kinesthetic and visual) between the unaffected side and the affected side in individuals with complex regional pain syndrome and chronic limb pain, there is also a five-fold probability that no differences exist.

However, we could not confirm the similarity between groups for the total and visual mean subscores.

## DISCUSSION

In this study we observed high inter-individual variability, with dispersion around the group medians ranging from 15% to 25% for total explicit motor imagery scores. There were no significant differences between groups or between the affected and unaffected side. A secondary analysis showed similar explicit motor imagery abilities between groups and between the affected and unaffected side. The significance of these findings will be discussed below.

First, our study revealed significant variability in explicit mental imagery abilities between participants, consistent with the broader spectrum of explicit imagery vividness identified in the literature, ranging from aphantasia to hyperphantasia, as reported by Zeman (Zeman, 2024). The MIQ-RS was unable to discriminate between healthy and painful individuals, suggesting that it may not effectively capture explicit motor imagery at the group level. The observed heterogeneity between individuals in explicit motor imagery abilities does not reflect changes in these abilities over time, nor does it allow us to understand the dynamics, i.e. whether individuals suffering from chronic pain had the ability to engage in explicit mental imagery prior to the onset of chronic pain. However, our results show that patients with chronic pain exhibit greater variability in ability compared to healthy individuals, a pattern consistent with the theory of pain-sensorimotor interactions (Murray & Sessle, 2024). This theory highlights the influence of biological, psychological and social factors on changes in sensorimotor behaviour in patients with chronic pain. Similarly, our findings suggest that the heterogeneity observed in sensorimotor changes may also extend to explicit motor imagery abilities.

Secondly, the absence of differences between groups for total mean scores and visual mean subscores was not confirmed by our Bayesian Null Hypothesis Testing. This may suggest that factors such as age may differentially affect explicit motor imagery abilities. Indeed, performance on implicit motor imagery tasks has been shown to be influenced by age (Muto et al., 2022). However, for explicit motor imagery tasks, age appears to differentially affect performance, with younger individuals showing greater visual dominance while older individuals showing stronger kinesthetic abilities, as differences are observed across different task types (Saimpont et al., 2015; Subirats et al., 2018). Furthermore, the brain areas involved in the modalities of motor imagery tasks differ (Lotze & Moseley, 2022b). Implicit and explicit motor imagery tasks activate similar areas, particularly in the beta band frequency of electroencephalography, but implicit tasks are less spatially specific and more intense than explicit tasks, suggesting different ways of mobilising sensorimotor areas (Osuagwu & Vuckovic, 2014). However, individuals can perform hand laterality judgment tasks (explicit motor imagery) without using a motor imagery-based strategy (Mibu et al., 2020). Furthermore, kinesthetic tasks have shown greater brain activation and correlation with brain areas in fMRI than visual modalities (Confalonieri et al., 2012; Lee et al., 2019). These findings provide additional confidence in our results.

Third, our findings contrast with previous studies that have identified effects of chronic pain on explicit motor imagery abilities (La Touche et al., 2019). In such studies, participants with chronic LBP exhibited more catastrophizing than healthy controls, with previous research showing a significant interaction between motor imagery and levels of catastrophizing (Moseley et al., 2008). Furthermore, a pilot study showed that stress conditions affect implicit but not explicit motor imagery abilities in healthy individuals (Schlatter et al., 2020), and it is well documented that participants with chronic pain report higher levels of stress (Mills et al., 2019). Furthermore, cognitive factors have been shown to influence implicit motor imagery tasks (Pelletier et al., 2018). These results could explain our discrepancy by highlighting the importance of the interaction between psychological factors and motor imagery abilities.

Motor imagery tasks target the same brain networks as voluntary motor movements and motor imagery therapy has been shown to be effective in improving neuronal excitability and synaptic conductance in both healthy and pathological individuals (Bowering et al., 2013; Decety, 1996; Lotze & Moseley, 2022a; Ríos-León et al., 2024; Ruffino et al., 2017). Interestingly, CRPS participants show no change in motor planning when engaged in object affordance tasks (Ten Brink et al., 2024), suggesting that motor imagery may represent a more conscious experience of motor planning, as described by the perceptual-cognitive model (Hurst & Boe, 2022). This finding highlights the potential of explicit motor imagery as an entry point for rehabilitation aimed at improving motor performance and reducing pain. However, engaging in explicit motor imagery tasks can induce pain and sudomotor symptoms in patients with complex regional pain syndrome (CRPS), suggesting the need for a graduated approach to exposure even during motor imagery therapy sessions (Moseley et al., 2008).

Explicit imagery training is the second stage of Graded Motor Imagery (GMI) therapy, which consists of three phases: starting with implicit motor imagery tasks, progressing to explicit motor imagery tasks, and ending with mirror therapy. The sequence of these phases is thought to be important for the benefits of the therapy (Lotze & Moseley, 2022b; Moseley, 2005). Despite its effectiveness, GMI shows inconsistent results, which could be explained by the interindividual variability observed in our study and previously described by others (Méndez-Rebolledo et al., 2017; Smart et al., 2022). Individuals with lower explicit motor imagery abilities could exhibit hypoactivation, which could explain a form of motor disuse or dysfunction (Kantak et al., 2022; Punt et al., 2013). Consequently, assessment of motor imagery vividness using more inclusive tools than the MIQ-RS could allow tailoring of rehabilitation programmes to individual needs prior to mirror therapy. This personalised approach could effectively address the different subtypes of CRPS and improve recovery outcomes (Knudsen et al., 2023; Mangnus et al., 2023).

All in all, our results suggest that chronic pain affects cortical function and structure (Yang & Chang, 2019) differently depending on the processes involved in motor behaviour, the type of pain, the presence of psychological factors or individual behavioral strategies related to motor imagery. Our findings are consistent with the notion of reciprocal changes in motor behaviour and plasticity induced by chronic pain, as well as clinical recovery and plasticity induced by exercise (Hodges & Smeets, 2015; Kourosh-Arami & Komaki, 2023; Merkle et al., 2020).

Limitations include the non-random order of questionnaire completion and the aggregation of upper, lower and spinal motor imagery scores in the calculation of the MIQ-RS score, which may have masked specific limb scores. Furthermore, the lack of homogeneity of the subgroups in terms of physical activity level, education level, and pain duration may have biased our results, although all participants were inactive (physical activity less than 2.5 hours per week) and all pain groups were chronic (pain duration more than three months). Educational level appears to be a predictor of pain chronicity (Prego-Domínguez et al., 2021), and despite the lower educational level in the pain groups, there were no differences in motor imagery abilities, which may mitigate the recruitment bias in our study. Our study did not assess pain intensity, which could have revealed potential sources of bias in different subtypes of chronic pain patients (Knudsen et al., 2023).

Future research should pursue a deeper understanding of the efficacy mechanisms underlying different motor imagery training tasks (explicit, implicit, external, internal) in patients with complex regional pain syndrome (Diers, 2019), using more objective measures such as brain imaging in longitudinal research designs. In addition, exploring patient identification methods for personalised interventions, similar to those explored in chronic low back pain research (Simula et al., 2020), may help to identify individuals suitable for specific interventions (Mangnus et al., 2023).

## CONCLUSION

Individuals with CRPS and chronic limb pain showed high inter-individual variability in explicit motor imagery tasks, similar to that observed in healthy people, with preserved abilities between groups and sides. This supports the recommendation of mental imagery therapy to improve motor function and reduce pain in chronic pain patients.

## Data Availability

All data produced in the present work are contained in the manuscript

## ACKNOWLEDGMENTS

We thank the following people for their support and help: Marine Ourmet, Brigitte Laffont for regulatory documents, Willy Fagart, Dr. Anaïs Pages, Julie Bourdier, Romain Dolin, Shuan Banh, Kevin Jezequel and Sarah Kabani for editing the manuscript. The authors declare no conflicts of interest. The study data are available upon reasonable request to the corresponding author.

## AUTHOR CONTRIBUTIONS

**G. Cohen-Aknine** : Conceptualization, Methodology, Formal Analysis, Investigation, Writing-Original Draft **A Homs:** Resources, Writing-Review & Editing, Visualization **D. Mottet:** Methodology, Validation, Data Curation, Writing-Review & Editing, Visualization, Supervision, **T. Mura:** Methodology, Writing-Review & Editing **F. Jedryka:** Investigation, Resources, Writing-Review & Editing **A. Dupeyron:** Validation, Resources, Writing-Review & Editing, Visualization, Project Administration

## DATA AVAIBILITY

The datasets generated during and/or analyzed during the current study are available from the corresponding author on reasonable request.

## Supplementary Information

Details of the demographic characteristics and Table S2, S3, S4, S5 & S6 are provided in Appendix S1.

Details of statistical analysis are provided in Appendix S2 in html.

**Appendix S1.** Participants demographic characteristics.

Participants demographic characteristics
| GROUP | AGE | SEX | UPPER DOMINANT | LOWER DOMINANT | BMI | DIPLOMA | LEVEL OF PHYSICAL ACTIVITY | PAIN DURATION | PAIN SIDE | PATHOLOGY |
| --- | --- | --- | --- | --- | --- | --- | --- | --- | --- | --- |
| HEALTHY | 21-25 | Male | Right | right | 21 | Master's Degree | >1h30 of moderate to high level per week |  |  |  |
| HEALTHY | 26-30 | Male | Right | right | 18 | 2 years of study after A level | < 1h of moderate to high level per week |  |  |  |
| HEALTHY | 31-35 | Male | Right | left | 22 | 2 years of study after A level | >1h30 of moderate to high level per week |  |  |  |
| HEALTHY | 36-40 | Male | Right | right | 21 | Bachelor degree | >1h30 of moderate to high level per week |  |  |  |
| HEALTHY | 36-40 | Male | Right | right | 29 | Master's Degree | >1h30 of moderate to high level per week |  |  |  |
| HEALTHY | 36-40 | Male | Right | right | 22 | A Level / High School diploma or equivalent | >1h and <1h30 of moderate to high level per week |  |  |  |
| HEALTHY | 36-40 | Male | Right | left | 20 | 2 years of study after A level | >1h30 of moderate to high level per week |  |  |  |
| HEALTHY | 36-40 | Male | Right | right | 30 | Bachelor degree | < 1h of moderate to high level per week |  |  |  |
| HEALTHY | 46-50 | Male | Right | right | 23 | Master's Degree | < 1h of moderate to high level per week |  |  |  |
| HEALTHY | 51-55 | Male | Right | right | 30 | Ph.D. and higher | < 1h of moderate to high level per week |  |  |  |
| HEALTHY | 51-55 | Male | Right | left | 25 | 1 years of study after A level | >1h and <1h30 of moderate to high level per week |  |  |  |
| HEALTHY | 51-55 | Male | Right | right | 25 | Master's Degree | >1h30 of moderate to high level per week |  |  |  |
| HEALTHY | 56-60 | Male | Right | right | 27 | Ph.D. and higher | < 1h of moderate to high level per week |  |  |  |
| HEALTHY | 56-60 | Male | Right | left | 29 | Ph.D. and higher | >1h and <1h30 of moderate to high level per week |  |  |  |
| HEALTHY | 66-70 | Male | Right | left | 31 | Ph.D. and higher | >1h30 of moderate to high level per week |  |  |  |
| HEALTHY | 66-70 | Male | Right | right | 24 | 1 years of study after A level | >1h and <1h30 of moderate to high level per week |  |  |  |
| HEALTHY | 21-25 | Female | Right | right | 20 | 2 years of study after A level | < 1h of moderate to high level per week |  |  |  |
| HEALTHY | 31-35 | Female | Right | right | 20 | Ph.D. and higher | >1h and <1h30 of moderate to high level per week |  |  |  |
| HEALTHY | 31-35 | Female | Right | left | 39 | 1 years of study after A level | < 1h of moderate to high level per week |  |  |  |
| HEALTHY | 31-35 | Female | Right | left | 27 | 1 years of study after A level | >1h and <1h30 of moderate to high level per week |  |  |  |
| HEALTHY | 36-40 | Female | Left | right | 21 | Ph.D. and higher | >1h and <1h30 of moderate to high level per week |  |  |  |
| HEALTHY | 36-40 | Female | Right | right | 36 | Bachelor degree | >1h30 of moderate to high level per week |  |  |  |
| HEALTHY | 36-40 | Female | Right | right | 20 | Bachelor degree | >1h30 of moderate to high level per week |  |  |  |
| HEALTHY | 36-40 | Female | Right | right | 19 | Bachelor degree | < 1h of moderate to high level per week |  |  |  |
| HEALTHY | 36-40 | Female | Right | right | 29 | 2 years of study after A level | < 1h of moderate to high level per week |  |  |  |
| HEALTHY | 41-45 | Female | Right | right | 21 | Master's Degree | >1h and <1h30 of moderate to high level per week |  |  |  |
| HEALTHY | 41-45 | Female | Right | right | 24 | Master's Degree | < 1h of moderate to high level per week |  |  |  |
| HEALTHY | 46-50 | Female | Right | left | 21 | Bachelor degree | < 1h of moderate to high level per week |  |  |  |
| HEALTHY | 46-50 | Female | Right | right | 21 | 2 years of study after A level | >1h and <1h30 of moderate to high level per week |  |  |  |
| HEALTHY | 46-50 | Female | Right | right | 20 | 2 years of study after A level | >1h and <1h30 of moderate to high level per week |  |  |  |
| HEALTHY | 46-50 | Female | Right | right | 28 | 2 years of study after A level | >1h and <1h30 of moderate to high level per week |  |  |  |
| HEALTHY | 46-50 | Female | Right | right | 25 | Master's Degree | >1h30 of moderate to high level per week |  |  |  |
| HEALTHY | 46-50 | Female | Right | right | 23 | Master's Degree | < 1h of moderate to high level per week |  |  |  |
| HEALTHY | 46-50 | Female | Right | left | 23 | Master's Degree | >1h and <1h30 of moderate to high level per week |  |  |  |
| HEALTHY | 51-55 | Female | Right | right | 21 | Master's Degree | < 1h of moderate to high level per week |  |  |  |
| HEALTHY | 51-55 | Female | Right | left | 21 | Bachelor degree | >1h and <1h30 of moderate to high level per week |  |  |  |
| HEALTHY | 51-55 | Female | Left | left | 20 | Master's Degree | >1h and <1h30 of moderate to high level per week |  |  |  |
| HEALTHY | 51-55 | Female | Right | right | 24 | Bachelor degree | < 1h of moderate to high level per week |  |  |  |
| HEALTHY | 56-60 | Female | Right | left | 28 | 2 years of study after A level | < 1h of moderate to high level per week |  |  |  |
| HEALTHY | 61-65 | Female | Right | right | 28 | Bachelor degree | < 1h of moderate to high level per week |  |  |  |
| HEALTHY | 61-65 | Female | Right | right | 25 | A Level / High School diploma or equivalent | >1h and <1h30 of moderate to high level per week |  |  |  |
| HEALTHY | 61-65 | Female | Right | left | 23 | 1 years of study after A level | >1h and <1h30 of moderate to high level per week |  |  |  |
| HEALTHY | 61-65 | Female | Right | left | 21 | 2 years of study after A level | >1h30 of moderate to high level per week |  |  |  |
| CRPS | 31-35 | Male | Right | right | 40 | 2 years of study after A level | >1h and <1h30 of moderate to high level per week | 4 | Right Lower Limb |  |
| CRPS | 36-40 | Male | Right | left | 26 | A Level / High School diploma or equivalent | >1h and <1h30 of moderate to high level per week | 7 | Right Lower Limb |  |
| CRPS | 46-50 | Male | Right | right | 24 | 1 years of study after A level | < 1h of moderate to high level per week | 8 | Left Lower Limb |  |
| CRPS | 46-50 | Male | Right | right | 30 | 2 years of study after A level | < 1h of moderate to high level per week | 8 | Right Upper Limb |  |
| CRPS | 51-55 | Male | Right | right | 20 | A Level / High School diploma or equivalent | < 1h of moderate to high level per week | 6 | Right Lower Limb |  |
| CRPS | 56-60 | Male | Right | right | 29 | A Level / High School diploma or equivalent | < 1h of moderate to high level per week | 8 | Left Lower Limb |  |
| CRPS | 61-65 | Male | Right | right | 26 | A Level / High School diploma or equivalent | < 1h of moderate to high level per week | 19 | Left Lower Limb |  |
| CRPS | 61-65 | Male | Right | right | 28 | A Level / High School diploma or equivalent | < 1h of moderate to high level per week | 5 | Right Lower Limb |  |
| CRPS | 66-70 | Male | Right | left | 30 | 1 years of study after A level | < 1h of moderate to high level per week | 15 | Left Lower Limb |  |
| CRPS | 21-25 | Female | Left | left | 21 | A Level / High School diploma or equivalent | < 1h of moderate to high level per week | 13 | Left Lower Limb |  |
| CRPS | 26-30 | Female | Right | left | 19 | 1 years of study after A level | < 1h of moderate to high level per week | 1 | Left Lower Limb |  |
| CRPS | 31-35 | Female | Right | right | 23 | A Level / High School diploma or equivalent | < 1h of moderate to high level per week | 15 | Right Upper Limb |  |
| CRPS | 31-35 | Female | Right | right | 29 | 1 years of study after A level | < 1h of moderate to high level per week | 13 | Left Lower Limb |  |
| CRPS | 31-35 | Female | Right | right | 23 | 1 years of study after A level | < 1h of moderate to high level per week | 10 | Right Upper Limb |  |
| CRPS | 36-40 | Female | Right | right | 20 | A Level / High School diploma or equivalent | < 1h of moderate to high level per week | 21 | Right Lower Limb |  |
| CRPS | 36-40 | Female | Right | right | 24 | 1 years of study after A level | < 1h of moderate to high level per week | 7 | Right Lower Limb |  |
| CRPS | 41-45 | Female | Left | right | 24 | 1 years of study after A level | < 1h of moderate to high level per week | 12 | Left Lower Limb |  |
| CRPS | 41-45 | Female | Right | right | 30 | 1 years of study after A level | < 1h of moderate to high level per week | 6 | Right Upper Limb |  |
| CRPS | 46-50 | Female | Left | right | 23 | A Level / High School diploma or equivalent | < 1h of moderate to high level per week | 90 | Left Upper Limb |  |
| CRPS | 46-50 | Female | Right | right | 25 | A Level / High School diploma or equivalent | < 1h of moderate to high level per week | 32 | Left Upper Limb |  |
| CRPS | 46-50 | Female | Right | left | 30 | A Level / High School diploma or equivalent | < 1h of moderate to high level per week | 36 | Left Lower Limb |  |
| CRPS | 46-50 | Female | Right | right | 18 | A Level / High School diploma or equivalent | < 1h of moderate to high level per week | 10 | Right Upper Limb |  |
| CRPS | 51-55 | Female | Right | left | 26 | 1 years of study after A level | < 1h of moderate to high level per week | 11 | Left Lower Limb |  |
| CRPS | 51-55 | Female | Left | left | 35 | A Level / High School diploma or equivalent | < 1h of moderate to high level per week | 13 | Right Lower Limb |  |
| CRPS | 51-55 | Female | Right | right | 22 | Bachelor degree | >1h30 of moderate to high level per week | 3 | Right Upper Limb |  |
| CRPS | 51-55 | Female | Right | left | 30 | A Level / High School diploma or equivalent | < 1h of moderate to high level per week | 10 | Left Upper Limb |  |
| CRPS | 51-55 | Female | Right | right | 23 | A Level / High School diploma or equivalent | >1h and <1h30 of moderate to high level per week | 26 | Right Lower Limb |  |
| CRPS | 51-55 | Female | Left | left | 26 | 1 years of study after A level | < 1h of moderate to high level per week | 6 | Left Upper Limb |  |
| CRPS | 56-60 | Female | Right | right | 40 | A Level / High School diploma or equivalent | < 1h of moderate to high level per week | 6 | Right Lower Limb |  |
| CRPS | 56-60 | Female | Right | right | 27 | A Level / High School diploma or equivalent | < 1h of moderate to high level per week | 9 | Right Lower Limb |  |
| CRPS | 56-60 | Female | Right | right | 29 | Bachelor degree | < 1h of moderate to high level per week | 11 | Right Upper Limb |  |
| CRPS | 56-60 | Female | Right | right | 22 | 2 years of study after A level | >1h30 of moderate to high level per week | 30 | Left Lower Limb |  |
| CRPS | 61-65 | Female | Right | right | 19 | A Level / High School diploma or equivalent | < 1h of moderate to high level per week | 5 | Left Lower Limb |  |
| CRPS | 61-65 | Female | Right | right | 28 | A Level / High School diploma or equivalent | < 1h of moderate to high level per week | 3 | Left Upper Limb |  |
| CRPS | 61-65 | Female | Right | right | 30 | A Level / High School diploma or equivalent | < 1h of moderate to high level per week | 8 | Left Lower Limb |  |
| CRPS | 61-65 | Female | Right | right | 25 | 1 years of study after A level | >1h and <1h30 of moderate to high level per week | 8 | Left Lower Limb |  |
| CRPS | 71-75 | Female | Right | right | 26 | A Level / High School diploma or equivalent | >1h30 of moderate to high level per week | 35 | Left Lower Limb |  |
| CRPS | 71-75 | Female | Right | right | 21 | A Level / High School diploma or equivalent | >1h30 of moderate to high level per week | 46 | Left Lower Limb |  |
| CRPS | 81-85 | Female | Right | right | 24 | A Level / High School diploma or equivalent | < 1h of moderate to high level per week | 7 | Left Upper Limb |  |
| CRPS | 81-85 | Female | Right | right | 23 | A Level / High School diploma or equivalent | < 1h of moderate to high level per week | 6 | Right Lower Limb |  |
| CLP | 18-21 | Male | Right | left | 18 | A Level / High School diploma or equivalent | >1h30 of moderate to high level per week | 5 | Right Lower Limb | Cruciate ligament surgery |
| CLP | 26-30 | Male | Right | right | 30 | Bachelor degree | >1h and <1h30 of moderate to high level per week | 2 | Left Upper Limb | Severe sprain |
| CLP | 31-35 | Male | Right | right | 30 | A Level / High School diploma or equivalent | >1h and <1h30 of moderate to high level per week | 54 | Right Lower Limb | menisectomy |
| CLP | 31-35 | Male | Left | left | 41 | Master's Degree | >1h30 of moderate to high level per week | 48 | Left Lower Limb | Arthrosis |
| CLP | 36-40 | Male | Right | right | 30 | A Level / High School diploma or equivalent | < 1h of moderate to high level per week | 1,5 | Right Upper Limb | Wrist pseudarthrosis |
| CLP | 41-45 | Male | Right | left | 48 | A Level / High School diploma or equivalent | < 1h of moderate to high level per week | 100 | Right Lower Limb | femoro acetabular conflict |
| CLP | 41-45 | Male | Right | left | 21 | Bachelor degree | >1h30 of moderate to high level per week | 26 | Left Upper Limb | Tendinopathy |
| CLP | 41-45 | Male | Right | right | 24 | A Level / High School diploma or equivalent | < 1h of moderate to high level per week | 8 | Right Upper Limb | Acromioclavicular disjunction |
| CLP | 46-50 | Male | Right | left | 26 | Master's Degree | < 1h of moderate to high level per week | 3 | Left Upper Limb | collarbone fracture |
| CLP | 46-50 | Male | Right | left | 28 | Ph.D. and higher | < 1h of moderate to high level per week | 60 | Left Lower Limb | osteonecrosis |
| CLP | 51-55 | Male | Right | right | 24 | 1 years of study after A level | < 1h of moderate to high level per week | 7 | Right Upper Limb | thumb ligamentoplasty D |
| CLP | 51-55 | Male | Right | left | 26 | 1 years of study after A level | < 1h of moderate to high level per week | 12 | Right Upper Limb | Carpal tunnel |
| CLP | 56-60 | Male | Right | left | 27 | 1 years of study after A level | >1h30 of moderate to high level per week | 35 | Right Upper Limb | epicondylitis |
| CLP | 66-70 | Male | Right | right | 30 | 1 years of study after A level | < 1h of moderate to high level per week | 60 | Right Lower Limb | Total Hip Replacement |
| CLP | 71-75 | Male | Right | right | 22 | A Level / High School diploma or equivalent | >1h30 of moderate to high level per week | 12 | Right Upper Limb | Tendinopathy |
| CLP | 76-80 | Male | Right | right | 28 | A Level / High School diploma or equivalent | >1h30 of moderate to high level per week | 0,5 | Left Lower Limb | Infection |
| CLP | 76-80 | Male | Right | left | 25 | A Level / High School diploma or equivalent | < 1h of moderate to high level per week | 18 | Left Lower Limb | Arthrosis |
| CLP | 26-30 | Female | Right | right | 24 | 2 years of study after A level | >1h30 of moderate to high level per week | 8 | Left Lower Limb | Meniscal tear |
| CLP | 26-30 | Female | Right | left | 31 | Master's Degree | >1h30 of moderate to high level per week | 37 | Right Upper Limb | Wrist fracture |
| CLP | 31-35 | Female | Right | right | 20 | 2 years of study after A level | < 1h of moderate to high level per week | 12 | Right Lower Limb | Ankle instability |
| CLP | 36-40 | Female | Right | right | 19 | Master's Degree | >1h30 of moderate to high level per week | 12 | Right Lower Limb | Femoro patellar syndrome |
| CLP | 36-40 | Female | Right | right | 24 | A Level / High School diploma or equivalent | >1h and <1h30 of moderate to high level per week | 72 | Left Upper Limb | Tendinopathy |
| CLP | 41-45 | Female | Right | right | 25 | 2 years of study after A level | >1h and <1h30 of moderate to high level per week | 12 | Right Upper Limb | Tendinopathy |
| CLP | 46-50 | Female | Right | right | 18 | A Level / High School diploma or equivalent | >1h30 of moderate to high level per week | 3 | Right Lower Limb | leg fracture and cruciate ligament tear |
| CLP | 46-50 | Female | Right | right | 24 | Bachelor degree | >1h and <1h30 of moderate to high level per week | 6 | Left Lower Limb | intra synovial kyste |
| CLP | 46-50 | Female | Right | right | 32 | Master's Degree | >1h30 of moderate to high level per week | 10 | Right Upper Limb | Tendinitis |
| CLP | 46-50 | Female | Right | right | 39 | Master's Degree | < 1h of moderate to high level per week | 5 | Right Lower Limb | malleolus fracture and sprain |
| CLP | 46-50 | Female | Right | left | 22 | A Level / High School diploma or equivalent | < 1h of moderate to high level per week | 15 | Left Upper Limb | Rotator cuff tendinopathy |
| CLP | 46-50 | Female | Right | right | 41 | Master's Degree | >1h30 of moderate to high level per week | 100 | Right Upper Limb | Tendinopathy |
| CLP | 51-55 | Female | Right | right | 22 | 1 years of study after A level | < 1h of moderate to high level per week | 7 | Left Upper Limb | Finger drop |
| CLP | 51-55 | Female | Right | right | 27 | Ph.D. and higher | >1h and <1h30 of moderate to high level per week | 48 | Left Upper Limb | Rhizarthrosis |
| CLP | 56-60 | Female | Right | right | 20 | A Level / High School diploma or equivalent | < 1h of moderate to high level per week | 60 | Left Lower Limb | Arthroscopy |
| CLP | 56-60 | Female | Right | right | 24 | 2 years of study after A level | >1h30 of moderate to high level per week | 8 | Left Upper Limb | Fracture |
| CLP | 56-60 | Female | Right | left | 23 | Ph.D. and higher | >1h30 of moderate to high level per week | 1 | Right Upper Limb | wrist fracture and disjunction |
| CLP | 61-65 | Female | Right | left | 26 | A Level / High School diploma or equivalent | < 1h of moderate to high level per week | 12 | Left Lower Limb | Total Knee Replacement |
| CLP | 61-65 | Female | Right | right | 29 | 2 years of study after A level | >1h30 of moderate to high level per week | 10 | Right Upper Limb | Carpal tunnel |
| CLP | 66-70 | Female | Right | left | 22 | Bachelor degree | < 1h of moderate to high level per week | 100 | Left Lower Limb | Knee arthrosis |
| CLP | 71-75 | Female | Right | right | 22 | Bachelor degree | >1h and <1h30 of moderate to high level per week | 2,5 | Right Lower Limb | Bimalleolar fracture |
| CLP | 71-75 | Female | Right | right | 22 | Bachelor degree | >1h30 of moderate to high level per week | 5 | Right Upper Limb | Rotator cuff tendinopathy |
| CLP | 81-85 | Female | Right | right | 39 | Bachelor degree | < 1h of moderate to high level per week | 24 | Right Lower Limb | Arthrosis |

**Table S2:** Total MIQ-RS scores for complex regional pain syndrome, chronic limb pain and healthy groups, side by side. Data are presented as median and interquartile range (IQR).

| **Groups**<br>*Median [IQR]* | **Total MIQ-RS score :**<br>**affected side** | **Total MIQ-RS score :**<br>**unaffected side** |
| --- | --- | --- |
| **CRPS** | 74 [24] | 71 [21.25] |
| **CLP** | 77.5 [16.25] | 74 [20.25] |
|  | **Total MIQ-RS score :**<br>**right side** | **Total MIQ-RS score :**<br>**left side** |
| **Healthy** | 76 [18.5] | 72 [21] |

**Table S3:** MIQ-RS mean scores for complex regional pain syndrome, chronic limb pain and healthy group. Data are presented as median and interquartile range (IQR).

| **Groups**<br>*Median [IQR]* | **CLP** | **CRPS** | **Healthy** | **P value** |
| --- | --- | --- | --- | --- |
| **MIQ-RS**<br>**Total Mean** | 5.54 [1.16]<br>(4.84-6) | 5.29 [1.71]<br>(4.29-6) | 5.36 [1.32]<br>(4.75-6.1) | 0.408 |
| **MIQ-RS**<br>**KMI Mean** | 5.21 [1.36]<br>(4.64-6) | 5.01 [2.04]<br>(3.86-5.89) | 5.29 [2.14]<br>(3.86-6) | 0.626 |
| **MIQ-RS**<br>**VMI Mean** | 5.93 [1.18]<br>(5.29-6.46) | 5.5 [1.39]<br>(4.64-6.04) | 6 [1.5]<br>(5-6.5) | 0.124 |

**Table S4:**
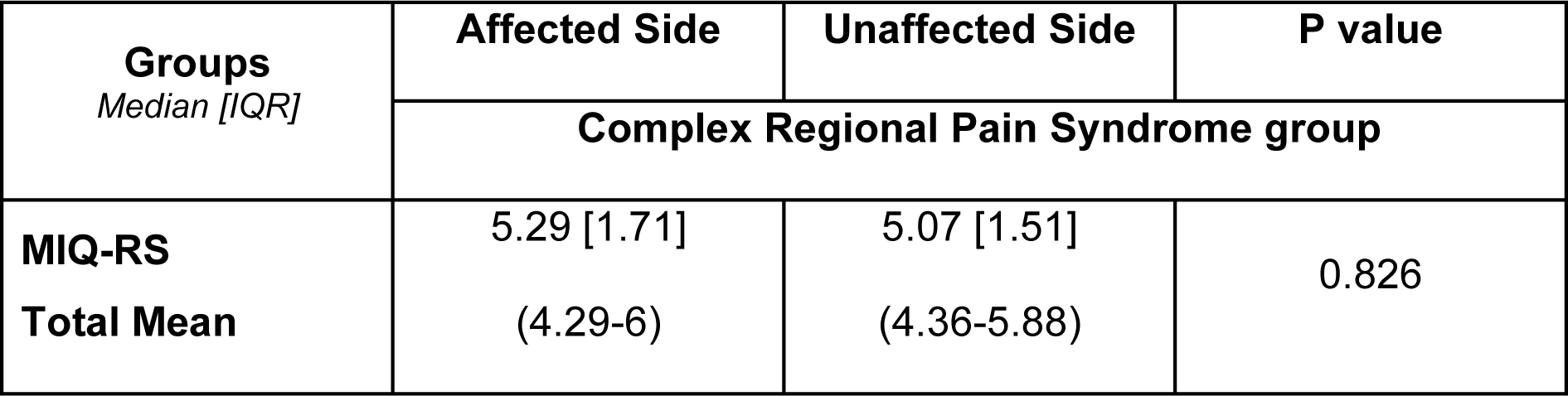
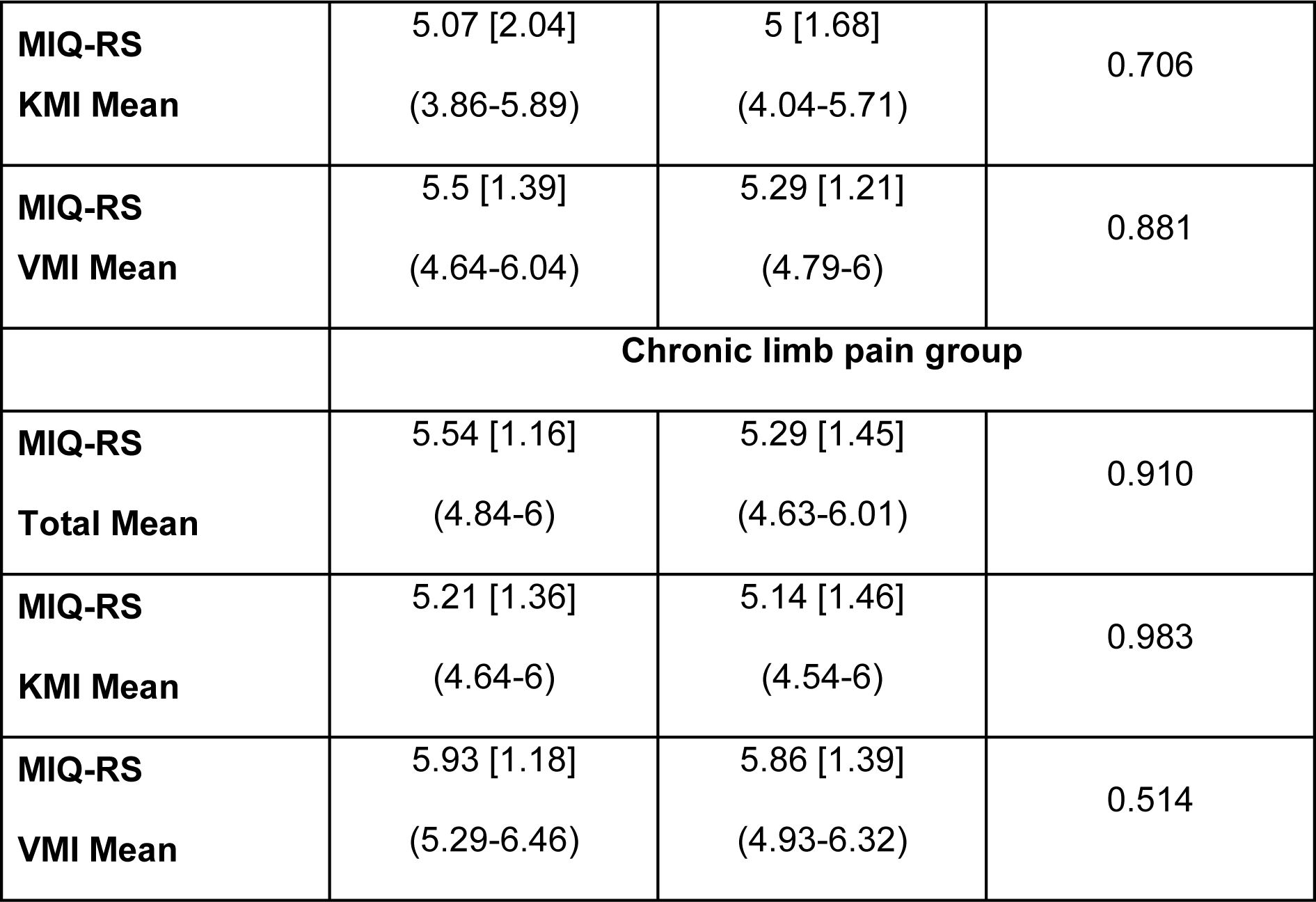
MIQ-RS mean subscores between the affected and unaffected side in patients with chronic limb pain (CLP). Data are presented as median and interquartile range (IQR).

**Table S5:** MIQ-RS mean subscores between the CRPS, CLP and healthy groups. Scores are expressed as Bayesian Factor BF01 according to the Bayesian Null Hypothesis Testing.

|  |  |
| --- | --- |
| (* $BF_{01} > 5$) | **BF<sub>01</sub>** |
| **MIQ-RS**<br>**Total Mean** | 2.57 |
| **MIQ-RS**<br>**KMI Mean** | **7.28\*** |
| **MIQ-RS**<br>**VMI Mean** | 1.07 |

**Table S6:** MIQ-RS mean subscores between the unaffected and affected side for the CRPS and CLP groups. Scores are expressed as Bayesian Factor BF01 according to the Bayesian Null Hypothesis Testing.

| *(\* BF<sub>01</sub> > 5)* | **BF<sub>01</sub>**<br>**CRPS Group** | **BF<sub>01</sub>**<br>**CLP Group** |
| --- | --- | --- |
| **MIQ-RS**<br>**Total Mean** | **6.57\*** | **6.65\*** |
| **MIQ-RS**<br>**KMI Mean** | **7.57\*** | **6.74\*** |
| **MIQ-RS**<br>**VMI Mean** | **5.84\*** | **9.86\*** |

## REFERENCES

1. Birklein, F., Ajit, S. K., Goebel, A., Perez, R. S. G. M., & Sommer, C. (2018). Complex regional pain syndrome—Phenotypic characteristics and potential biomarkers. Nature Reviews Neurology, 14(5), 272–284. 10.1038/nrneurol.2018.20

2. Bowering, K. J., O’Connell, N. E., Tabor, A., Catley, M. J., Leake, H. B., Moseley, G. L., & Stanton, T. R. (2013). The Effects of Graded Motor Imagery and Its Components on Chronic Pain : A Systematic Review and Meta-Analysis. The Journal of Pain, 14(1), 3–13. 10.1016/j.jpain.2012.09.007

3. Breckenridge, J. D., Ginn, K. A., Wallwork, S. B., & McAuley, J. H. (2019). Do People With Chronic Musculoskeletal Pain Have Impaired Motor Imagery? A Meta-analytical Systematic Review of the Left/Right Judgment Task. The Journal of Pain, 20(2), 119–132. 10.1016/j.jpain.2018.07.004

4. Butler, A. J., Cazeaux, J., Fidler, A., Jansen, J., Lefkove, N., Gregg, M., Hall, C., Easley, K. A., Shenvi, N., & Wolf, S. L. (2012). The Movement Imagery Questionnaire-Revised, Second Edition (MIQ-RS) Is a Reliable and Valid Tool for Evaluating Motor Imagery in Stroke Populations. Evidence-Based Complementary and Alternative Medicine, 2012, 1–11. 10.1155/2012/497289

5. Chang, W.-J., O’Connell, N. E., Beckenkamp, P. R., Alhassani, G., Liston, M. B., & Schabrun, S. M. (2018). Altered Primary Motor Cortex Structure, Organization, and Function in Chronic Pain : A Systematic Review and Meta-Analysis. The Journal of Pain, 19(4), 341–359. 10.1016/j.jpain.2017.10.007

6. Chepurova, A., Hramov, A., & Kurkin, S. (2022). Motor Imagery : How to Assess, Improve Its Performance, and Apply It for Psychosis Diagnostics. Diagnostics, 12(4), 949. 10.3390/diagnostics12040949

7. Confalonieri, L., Pagnoni, G., Barsalou, L. W., Rajendra, J., Eickhoff, S. B., & Butler, A. J. (2012). Brain Activation in Primary Motor and Somatosensory Cortices during Motor Imagery Correlates with Motor Imagery Ability in Stroke Patients. ISRN Neurology, 2012, 613595. 10.5402/2012/613595

8. Debarnot, U., Sperduti, M., Di Rienzo, F., & Guillot, A. (2014). Experts bodies, experts minds : How physical and mental training shape the brain. Frontiers in Human Neuroscience, 8. https://www.frontiersin.org/article/10.3389/fnhum.2014.00280

9. Decety, J. (1996). Do imagined and executed actions share the same neural substrate? Cognitive Brain Research, 3(2), 87–93. 10.1016/0926-6410(95)00033-X

10. Diers, M. (2019). Neuroimaging the pain network—Implications for treatment. Best Practice & Research. Clinical Rheumatology, 33(3), 101418. 10.1016/j.berh.2019.05.003

11. Ferraro, M. C., O’Connell, N. E., Sommer, C., Goebel, A., Bultitude, J. H., Cashin, A. G., Moseley, G. L., & McAuley, J. H. (2024). Complex regional pain syndrome : Advances in epidemiology, pathophysiology, diagnosis, and treatment. The Lancet. Neurology, 23(5), 522–533. 10.1016/S1474-4422(24)00076-0

12. Ferraro, M., Cashin, A., Wand, B., Smart, K., Berryman, C., Marston, L., Moseley, G., McAuley, J., & O’Connell, N. (2023). Interventions for treating pain and disability in adults with complex regional pain syndrome-an overview of systematic reviews. Cochrane Database of Systematic Reviews, 6. 10.1002/14651858.CD009416.pub3

13. Floridou, G. A., Peerdeman, K. J., & Schaefer, R. S. (2022). Individual differences in mental imagery in different modalities and levels of intentionality. Memory & Cognition, 50(1), 29–44. 10.3758/s13421-021-01209-7

14. Goebel, A., Barker, C., Birklein, F., Brunner, F., Casale, R., Eccleston, C., Eisenberg, E., McCabe, C. S., Moseley, G. L., Perez, R., Perrot, S., Terkelsen, A., Thomassen, I., Zyluk, A., & Wells, C. (2019). Standards for the diagnosis and management of complex regional pain syndrome : Results of a European Pain Federation task force. European Journal of Pain, 23(4), 641–651. 10.1002/ejp.1362

15. Goebel, A., Birklein, F., Brunner, F., Clark, J. D., Gierthmühlen, J., Harden, N., Huygen, F., Knudsen, L., McCabe, C., Lewis, J., Maihöfner, C., Magerl, W., Moseley, G. L., Terkelsen, A., Thomassen, I., & Bruehl, S. (2021). The Valencia consensus-based adaptation of the IASP complex regional pain syndrome diagnostic criteria. Pain, 162(9), 2346–2348. 10.1097/j.pain.0000000000002245

16. Gregg, M., Hall, C., & Butler, A. (2010). The MIQ-RS : A Suitable Option for Examining Movement Imagery Ability. Evidence-Based Complementary and Alternative Medicine, 7(2), 249–257. 10.1093/ecam/nem170

17. Guillot, A., & Collet, C. (2005). Contribution from neurophysiological and psychological methods to the study of motor imagery. Brain Research. Brain Research Reviews, 50(2), 387–397. 10.1016/j.brainresrev.2005.09.004

18. Halicka, M., Vittersø, A. D., Proulx, M. J., & Bultitude, J. H. (2020). Neuropsychological Changes in Complex Regional Pain Syndrome (CRPS). Behavioural Neurology, 2020, e4561831. 10.1155/2020/4561831

19. Harden, R. N., Bruehl, S., Perez, R. S. G. M., Birklein, F., Marinus, J., Maihofner, C., Lubenow, T., Buvanendran, A., Mackey, S., Graciosa, J., Mogilevski, M., Ramsden, C., Chont, M., & Vatine, J.-J. (2010). Validation of proposed diagnostic criteria (the “Budapest Criteria”) for Complex Regional Pain Syndrome. Pain, 150(2), 268–274. 10.1016/j.pain.2010.04.030

20. Harden, R. N., McCabe, C. S., Goebel, A., Massey, M., Suvar, T., Grieve, S., & Bruehl, S. (2022). Complex Regional Pain Syndrome : Practical Diagnostic and Treatment Guidelines, 5th Edition. Pain Medicine, 23(Supplement_1), S1–S53. 10.1093/pm/pnac046

21. Hardwick, R. M., Caspers, S., Eickhoff, S. B., & Swinnen, S. P. (2018). Neural correlates of action : Comparing meta-analyses of imagery, observation, and execution. Neuroscience and Biobehavioral Reviews, 94, 31–44. 10.1016/j.neubiorev.2018.08.003

22. Hurst, A. J., & Boe, S. G. (2022). Imagining the way forward : A review of contemporary motor imagery theory. Frontiers in Human Neuroscience, 16. 10.3389/fnhum.2022.1033493

23. Iwatsuki, K., Hoshiyama, M., Yoshida, A., Uemura, J.-I., Hoshino, A., Morikawa, I., Nakagawa, Y., & Hirata, H. (2021). Chronic pain-related cortical neural activity in patients with complex regional pain syndrome. IBRO Neuroscience Reports, 10, 208–215. 10.1016/j.ibneur.2021.05.001

24. Kim, H., Lee, C.-H., Kim, S.-H., & Kim, Y.-D. (2018). Epidemiology of complex regional pain syndrome in Korea : An electronic population health data study. PloS One, 13(6), e0198147. 10.1371/journal.pone.0198147

25. Knudsen, L. F., Terkelsen, A. J., Drummond, P. D., & Birklein, F. (2019). Complex regional pain syndrome : A focus on the autonomic nervous system. Clinical Autonomic Research, 29(4), 457–467. 10.1007/s10286-019-00612-0

26. Kosek, E., Clauw, D., Nijs, J., Baron, R., Gilron, I., Harris, R. E., Mico, J.-A., Rice, A. S. C., & Sterling, M. (2021). Chronic nociplastic pain affecting the musculoskeletal system : Clinical criteria and grading system. PAIN, 162(11), 2629. 10.1097/j.pain.0000000000002324

27. Kruschke, J. K. (2021). Bayesian Analysis Reporting Guidelines. Nature Human Behaviour, 5(10), Article 10. 10.1038/s41562-021-01177-7

28. La Touche, R., Grande-Alonso, M., Cuenca-Martínez, F., Gónzález-Ferrero, L., Suso-Martí, L., & Paris-Alemany, A. (2019). Diminished Kinesthetic and Visual Motor Imagery Ability in Adults With Chronic Low Back Pain. *PM & R: The Journal of Injury*, Function, and Rehabilitation, 11(3), 227–235. 10.1016/j.pmrj.2018.05.025

29. Loison, B., Moussaddaq, A.-S., Cormier, J., Richard, I., Ferrapie, A.-L., Ramond, A., & Dinomais, M. (2013). Translation and validation of the French Movement Imagery Questionnaire – Revised Second version (MIQ-RS). Annals of Physical and Rehabilitation Medicine, 56(3), 157–173. 10.1016/j.rehab.2013.01.001

30. Lotze, M., & Moseley, G. L. (2022). Clinical and Neurophysiological Effects of Progressive Movement Imagery Training for Pathological Pain. The Journal of Pain, 23(9), 1480–1491. 10.1016/j.jpain.2022.04.008

31. McInnes, K., Friesen, C., & Boe, S. (2016). Specific Brain Lesions Impair Explicit Motor Imagery Ability : A Systematic Review of the Evidence. Archives of Physical Medicine and Rehabilitation, 97(3), 478–489.e1. 10.1016/j.apmr.2015.07.012

32. Mesaroli, G., Hundert, A., Birnie, K. A., Campbell, F., & Stinson, J. (2021). Screening and diagnostic tools for complex regional pain syndrome : A systematic review. PAIN, 162(5), 1295–1304. 10.1097/j.pain.0000000000002146

33. Mohan, V., Bhat, A., & Morasso, P. (2019). Muscleless motor synergies and actions without movements : From motor neuroscience to cognitive robotics. Physics of Life Reviews, 30, 89–111. 10.1016/j.plrev.2018.04.005

34. Moran, A., Guillot, A., MacIntyre, T., & Collet, C. (2012). Re-imagining motor imagery : Building bridges between cognitive neuroscience and sport psychology. British Journal of Psychology, 103(2), 224–247. 10.1111/j.2044-8295.2011.02068.x

35. Moseley, G. L. (2005). Is successful rehabilitation of complex regional pain syndrome due to sustained attention to the affected limb? A randomised clinical trial. Pain, 114(1), 54–61. 10.1016/j.pain.2004.11.024

36. Moseley, G. L., Zalucki, N., Birklein, F., Marinus, J., van Hilten, J. J., & Luomajoki, H. (2008). Thinking about movement hurts : The effect of motor imagery on pain and swelling in people with chronic arm pain. Arthritis & Rheumatism, 59(5), 623–631. 10.1002/art.23580

37. Muto, H., Suzuki, M., & Sekiyama, K. (2022). Advanced aging effects on implicit motor imagery and its links to motor performance : An investigation via mental rotation of letters, hands, and feet. Frontiers in Aging Neuroscience, 14. 10.3389/fnagi.2022.1025667

38. Ott, S., & Maihöfner, C. (2018a). Signs and Symptoms in 1,043 Patients with Complex Regional Pain Syndrome. The Journal of Pain, 19(6), 599–611. 10.1016/j.jpain.2018.01.004

39. Ott, S., & Maihöfner, C. (2018b). Signs and Symptoms in 1,043 Patients with Complex Regional Pain Syndrome. The Journal of Pain, 19(6), 599–611. 10.1016/j.jpain.2018.01.004

40. Petersen, P. B., Mikkelsen, K. L., Lauritzen, J. B., & Krogsgaard, M. R. (2018). Risk Factors for Post-treatment Complex Regional Pain Syndrome (CRPS) : An Analysis of 647 Cases of CRPS from the Danish Patient Compensation Association. Pain Practice, 18(3), 341–349. 10.1111/papr.12610

41. Prego-Domínguez, J., Khazaeipour, Z., Mallah, N., & Takkouche, B. (2021). Socioeconomic status and occurrence of chronic pain : A meta-analysis. Rheumatology, 60(3), 1091–1105. 10.1093/rheumatology/keaa758

42. Punt, D. T., Cooper, L., Hey, M., & Johnson, M. I. (2013). Neglect-like symptoms in complex regional pain syndrome : Learned nonuse by another name? PAIN, 154(2), 200. 10.1016/j.pain.2012.11.006

43. Quintana, D. S., & Williams, D. R. (2018). Bayesian alternatives for common null-hypothesis significance tests in psychiatry : A non-technical guide using JASP. BMC Psychiatry, 18(1), 178. 10.1186/s12888-018-1761-4

44. Ravat, S., Olivier, B., Gillion, N., & Lewis, F. (2020). Laterality judgment performance between people with chronic pain and pain-free individuals. A systematic review and meta-analysis. Physiotherapy Theory and Practice, 36(12), 1279–1299. 10.1080/09593985.2019.1570575

45. Rimbert, S., Gayraud, N., Bougrain, L., Clerc, M., & Fleck, S. (2019). Can a Subjective Questionnaire Be Used as Brain-Computer Interface Performance Predictor? Frontiers in Human Neuroscience, 12, 529. 10.3389/fnhum.2018.00529

46. Ríos-León, M., Cuñado-González, Á., Domínguez-Fernández, S., & Martín-Casas, P. (2024). Effectiveness of motor imagery in complex regional pain syndrome : A systematic review with meta-analysis. Pain Practice: The Official Journal of World Institute of Pain. 10.1111/papr.13348

47. Ruffino, C., Papaxanthis, C., & Lebon, F. (2017). Neural plasticity during motor learning with motor imagery practice : Review and perspectives. Neuroscience, 341, 61–78. 10.1016/j.neuroscience.2016.11.023

48. Saimpont, A., Malouin, F., Tousignant, B., & Jackson, P. L. (2015). Assessing motor imagery ability in younger and older adults by combining measures of vividness, controllability and timing of motor imagery. Brain Research, 1597, 196–209. 10.1016/j.brainres.2014.11.050

49. Schmidt, H., Drusko, A., Renz, M., Schlömp, L., Tost, H., Tesarz, J., Schuh-Hofer, S., Meyer-Lindenberg, A., & Treede, R.-D. (2023). Application of the IASP grading system for ‘nociplastic pain’ in chronic pain conditions : A field study (p. 2022.12.06.22283114). medRxiv. 10.1101/2022.12.06.22283114

50. Seiler, B. D., Newman-Norlund, R. D., & Monsma, E. V. (2017). Inter-individual neural differences in movement imagery abilities. Psychology of Sport and Exercise, 30, 153–163. 10.1016/j.psychsport.2017.02.007

51. Shafiee, E., MacDermid, J., Packham, T., Grewal, R., Farzad, M., Bobos, P., & Walton, D. (2023). Rehabilitation Interventions for Complex Regional Pain Syndrome : An Overview of Systematic Reviews. The Clinical Journal of Pain, 39(9), 473–483. 10.1097/AJP.0000000000001133

52. Sharma, N., Jones, P. S., Carpenter, T. A., & Baron, J.-C. (2008). Mapping the involvement of BA 4a and 4p during Motor Imagery. NeuroImage, 41(1), 92–99. 10.1016/j.neuroimage.2008.02.009

53. Shokouhi, M., Clarke, C., Morley-Forster, P., Moulin, D. E., Davis, K. D., & St Lawrence, K. (2018). Structural and Functional Brain Changes at Early and Late Stages of Complex Regional Pain Syndrome. The Journal of Pain, 19(2), 146–157. 10.1016/j.jpain.2017.09.007

54. Simula, A. S., Malmivaara, A., Booth, N., & Karppinen, J. (2020). A classification-based approach to low back pain in primary care—Protocol for a benchmarking controlled trial. BMC Family Practice, 21(1), 61. 10.1186/s12875-020-01135-8

55. Smart, K. M., Ferraro, M. C., Wand, B. M., & O’Connell, N. E. (2022). Physiotherapy for pain and disability in adults with complex regional pain syndrome (CRPS) types I and II. Cochrane Database of Systematic Reviews, 5. 10.1002/14651858.CD010853.pub3

56. Solomon, J. P., Hurst, A. J., Lee, J., & Boe, S. G. (2022). Are observed effects of movement simulated during motor imagery performance? Behavioral Neuroscience, 136(3), 264–275. 10.1037/bne0000517

57. Subirats, L., Allali, G., Briansoulet, M., Salle, J. Y., & Perrochon, A. (2018). Age and gender differences in motor imagery. Journal of the Neurological Sciences, 391, 114–117. 10.1016/j.jns.2018.06.015

58. Swart, C. M. A. K., Stins, J. F., & Beek, P. J. (2009). Cortical changes in complex regional pain syndrome (CRPS). *European Journal of Pain (London*, England), 13(9), 902–907. 10.1016/j.ejpain.2008.11.010

59. Ten Brink, A. F., & Bultitude, J. H. (2021). Predictors of Self-Reported Neglect-like Symptoms and Involuntary Movements in Complex Regional Pain Syndrome Compared to Other Chronic Limb Pain Conditions. Pain Medicine (Malden, Mass.), 22(10), 2337–2349. 10.1093/pm/pnab226

60. van der Meulen, M., Allali, G., Rieger, S. W., Assal, F., & Vuilleumier, P. (2014). The influence of individual motor imagery ability on cerebral recruitment during gait imagery. Human Brain Mapping, 35(2), 455–470. 10.1002/hbm.22192

61. van Doorn, J., van den Bergh, D., Böhm, U., Dablander, F., Derks, K., Draws, T., Etz, A., Evans, N. J., Gronau, Q. F., Haaf, J. M., Hinne, M., Kucharský, Š., Ly, A., Marsman, M., Matzke, D., Gupta, A. R. K. N., Sarafoglou, A., Stefan, A., Voelkel, J. G., & Wagenmakers, E.-J. (2021). The JASP guidelines for conducting and reporting a Bayesian analysis. Psychonomic Bulletin & Review, 28(3), 813–826. 10.3758/s13423-020-01798-5

62. van Velzen, G. A. J., Marinus, J., van Dijk, J. G., van Zwet, E. W., Schipper, I. B., & van Hilten, J. J. (2015). Motor Cortical Activity During Motor Tasks Is Normal in Patients With Complex Regional Pain Syndrome. The Journal of Pain, 16(1), 87–94. 10.1016/j.jpain.2014.10.010

63. Yang, S., & Chang, M. C. (2019). Chronic Pain : Structural and Functional Changes in Brain Structures and Associated Negative Affective States. International Journal of Molecular Sciences, 20(13), Article 13. 10.3390/ijms20133130

64. Yap, B. W. D., & Lim, E. C. W. (2019). The Effects of Motor Imagery on Pain and Range of Motion in Musculoskeletal Disorders : A Systematic Review Using Meta-Analysis. The Clinical Journal of Pain, 35(1), 87–99. 10.1097/AJP.0000000000000648

65. Zangrandi, A., Allen Demers, F., & Schneider, C. (2021). Complex Regional Pain Syndrome. A Comprehensive Review on Neuroplastic Changes Supporting the Use of Non-invasive Neurostimulation in Clinical Settings. Frontiers in Pain Research, 2. https://www.frontiersin.org/article/10.3389/fpain.2021.732343

